# Improving the health of people experiencing homelessness with recent drug overdose: rationale for and design of the Pharmacist and Homeless Outreach worker Engagement Non-medical Independent prescribing Rx (PHOENIx) pilot randomised controlled trial

**DOI:** 10.1101/2022.05.16.22275160

**Authors:** Richard Lowrie, Andrew McPherson, Frances S Mair, Donogh Maguire, Vibhu Paudyal, Becky Blair, David Brannan, Jane Moir, Fiona Hughes, Clare Duncan, Kate Stock, Natalia Farmer, Becci Ramage, Cian Lombard, Steven Ross, Ailsa Scott, George Provan, Laura Sills, Jennifer Hislop, Andrea Williamson

## Abstract

**Introduction:** Numbers of People Experiencing Homelessness (PEH) are increasing worldwide. Systematic reviews show high levels of multimorbidity and mortality due to treatable diseases including drug overdose. Integrated health and social care outreach interventions may improve outcomes. No previous studies have targeted PEH with recent drug overdose despite their high recorded levels of drug related deaths. There are few data on health and social care problems. Feasibility work suggests a collaborative health and social care intervention (Pharmacist and Homeless Outreach Engagement Non-medical Independent prescriber Rx-PHOENIx) is potentially beneficial. We describe the methods of a pilot randomised controlled trial(RCT) with parallel process and economic evaluation in PEH with at least one drug-related overdose in the preceding 6 months.

**Methods:** Detailed health and social care information will be collected at baseline before 1:1 randomisation to: care-as-usual plus visits from a pharmacist and homeless outreach worker (PHOENIx) for 6-9 months; or care-as-usual. The main outcomes are rate of presentations to emergency department(ED) for overdose or other causes and whether to progress to a definitive RCT based on: recruitment of ≥ 100 participants within 4 months;≥ 60% patients remaining in the study at 6 and 9 months follow up;≥ 60% of patients in the PHOENIx group receiving the intervention; and≥ 80% of patients with data collected. Secondary outcomes include: hospitalisations; treatment uptake and patient reported measures. Semi-structured interviews will explore future implementation of PHOENIx, and reasons for overdose and protective factors. An economic evaluation will assess the feasibility of conducting a cost effectiveness analysis in a subsequent definitive trial.

**Discussion:** The study will determine whether to proceed to a definitive RCT for PEH aiming to fulfil unmet health and social care needs of those experiencing homelessness and at risk of drug related harms and deaths while providing useful insights into barriers and facilitators to PHOENIx and characterising the health and social care needs of PEH.

**Ethics and dissemination:** The trial was approved by the South East Scotland National Health Service Research Ethics Committee 01. Results will be available in the last quarter of 2022.

**Registration details:** The trial is registered with the UK Clinical Trials Registry (ISRCTN 10585019).

**Strengths and limitations of the study:**

- We plan to recruit patients normally excluded from intervention trials and collect a diverse health and social care dataset at baseline
- The 6-9 month individualised, complex intervention offers longer consultations, integrated continuous, health and social care support on outreach
- Mixed methods will enable determination of whether a subsequent trial is merited from an efficacy, economic and patient perspective.
- By design, the pilot randomised controlled trial lacks the power to detect a clinically significant effect and recruitment was limited to 20 locations in Scotland’s largest city

## INTRODUCTION

### Background

Homelessness is a global problem, and the number of people experiencing homelessness (PEH) is increasing worldwide.^1^ The individual, societal, health and economic burden of homelessness is widely known and undisputed.^2^ The health of PEH is characterised by problems with mental health, physical health, drug and alcohol use and PEH tend to die prematurely aged 41-51 years, with numbers of long term conditions on a par with housed individuals almost twice their age.^3-6^ A majority of PEH experience problem poly drug use, and associated high rates of deaths through overdose.^7,8^ Other causes of death are also increasing.^5^ Homelessness is an independent risk factor for hospital admission and emergency department (ED) attendance and rates of ED use are increasing across healthcare systems worldwide.^2^ PEH are known to present to ED late and with comparatively more serious problems. ^2, 9^ PEH can be overwhelmed by their multiple health and social care problems and lack of support, experiencing individual, structural and institutional difficulties with self-care.^10-13^

The development and testing of complex, integrated health and social care interventions has been highlighted as an important priority with the associated need to test the impact of longer contact times.^2, 4, 5, 14, 16^

### Complex interventions for multiple problems

Assertive outreach delivered by workers who can establish trust and develop positive interpersonal relationships is an evidenced approach to strengthening primary health and social care for PEH. ^14-17^ Partnerships should be built between outreach health services and homeless service providers who are best placed to support wider needs including housing, education and employment.^14^ PEH perspectives on effective components of interventions include: involvement of peer workers in outreach programmes; help with housing; welfare payments and social prescribing. ^9, 14, 18, 19^ Interventions limited to addressing single health problems e.g. problem drug use, or mental health problems, may hold little appeal for PEH who have multiple problems. ^2, 15^

Interventions in published studies offered housing improvements for PEH with mental health or substance misuse, or target and address single physical health conditions e.g. HIV or Tuberculosis without addressing multimorbidity ^16, 19-23^ or diseases thought to be amenable to early intervention e.g. cardiovascular or respiratory disease.^5^ Of the few robust studies of interventions led by healthcare professionals aiming to improve broad outcomes e.g. mortality or reduced ED utilisation, none have been found to be effective. ^23^ Interventions to address multiple needs in PEH are necessarily complex, with a new framework supporting rigorous, phased testing.^24^ We are not aware of studies having following the recommended development stages for testing of complex interventions, including pilot testing to inform power and sample size calculation for a subsequent definitive trial. ^19, 24, 25^ There are no UK based intervention studies targeting community based PEH. ^23^

### A role for pharmacists collaborating with third sector homeless workers

Generalists may be best equipped to address the diverse levels of multimorbidity experienced by PEH, ^4, 15, 16^ suggesting the testing of holistic medical plus social care on outreach for community dwelling PEH is overdue. However, as workforce shortages worsened by COVID-19 may limit expansion of roles of established primary care clinicians, there may be a role for pharmacists with generalist independent clinical prescribing qualifications. Generalist Pharmacist independent prescribers exist across the world, offering a potential solution to ongoing General Practitioner and Nurse workforce shortages.^26^ Over 7500 (13%) of UK based pharmacists have undergone additional subsequent training in therapeutics and completed a period of additional supervised clinical training, to gain an independent prescribing qualification. This independent prescribing qualification enables diagnosis and prescribing for any condition within the pharmacist’s competency. In the UK, Pharmacists practise as part of the multidisciplinary health team throughout primary care (in community pharmacies as independent contractors or based in General Practice offices as clinical pharmacists) and secondary care. A collaboration between Pharmacists and third sector homeless workers is likely to be welcomed by PEH. ^10^ Emerging evidence of under-treatment with medicines ^3, 6^ and challenges to medicine adherence, ^27^ which may be amenable to pharmacist intervention ^28, 29^ both of which may contribute to poor health and premature death in PEH,^2, 5^ suggests a need to robustly test pharmacist led integrated health and social care intervention for PEH.

### PHOENIx

Staff working for homeless charities in Glasgow, Scotland (The Simon Community Scotland and The Marie Trust) have lived experience of homelessness and formed a novel, integrated National Health Service – Homeless sector partnership called PHOENIx**: P**harmacist and **H**omeless **O**utreach **E**ngagement and **N**on-medical **I**ndependent prescribing R**x**.^30-32^ The PHOENIx team assertively outreach to various locations in Glasgow e.g. homeless congregate accommodation, to engage and offer holistic assessment, treatment, prescribing and referral for patients’ expressed health and social care priorities. ^30-32^ PHOENIx is a secondary prevention intervention aiming to improve self-care and strengthen primary care to reduce the use of ED. The Pharmacist is from the National Health Service and the third sector outreach worker is from either of Glasgow’s homeless charities (Simon Community Scotland or the Marie Trust). Visiting patients once weekly, and with consultations lasting an hour on average, previous qualitative work suggests benefit to patients ^33^ and a feasibility study describes the pharmacist assessing, treating and prescribing for acute and chronic health problems, while the homeless charity link worker addresses benefits, housing and social prescribing.^32^ Working within the clinical governance framework provided by the patient’s GP and the local ED, PHOENIx may improve health and reduce emergency health service contacts. ^32, 33^

### PHOENIx after overdose pilot randomised controlled trial

Here, we describe methods from an ongoing pragmatic pilot randomised controlled multicentre trial with embedded economic and qualitative evaluation, of PHOENIx intervention targeting PEH with recent non-fatal drug overdose. The trial aim is to determine whether progression to a subsequent definitive randomised controlled trial is justified based on: reduction in ED visits; rate of participant recruitment and retention at 6 and 9 month follow up; fidelity of intervention delivery; and sufficient data collection at baseline and 6 and 9 month follow up. Given the paucity of data informing research and service delivery, we will also collect a diverse range of patient level health and social care data. Secondary outcome data include measures of overdose occurrence, healthcare utilisation, acceptability, and patient reported outcomes (e.g. quality of life/perceived burden of treatment/levels of anxiety/depression).

## METHODS

### Study setting

NHS Greater Glasgow and Clyde (GG&C) provides free primary, secondary and tertiary care to approximately 1.2 million people (almost 25% of the Scottish population). Prior to the COVID-19 pandemic, there were an estimated 7000 PEH in Glasgow every year with an estimated 560 rough sleeping annually, both proportions higher than in the rest of the UK.^30^ The study is set in 20 Glasgow venues (homeless accommodation or drop in centres).

### Eligibility criteria

#### Participants

Homeless individuals ^14^ aged 18 years and over are considered eligible if they have at least one drug overdose in the previous 6 months (Table 1).

**Table 1:** Trial Inclusion and Exclusion Criteria.

| Inclusion Criteria | Exclusion Criteria |
| --- | --- |
| Legally homeless (living in temporary homeless accommodation, rough sleeping or threatened with homelessness)<br>and<br>Aged 18 years and over | Living in residential or community based rehabilitation facility which has direct access to in-house medical and nursing care<br>or |
| and<br>One or more non prescribed drug overdoses in past six months confirmed by self-report and witnessed overdose/ambulance call out/ED visit /naloxone use | Unable to give written informed consent |

### PHOENIx team

The PHOENIx team comprise five part-time NHS Greater Glasgow and Clyde employee pharmacists working in pairs with four part-time Homeless charity link workers (from The Simon Community Scotland or The Marie Trust) available Monday to Friday, between the hours of 9am -6pm. Pharmacists are within the governance of a Homeless Health Service General Practice and a major teaching hospital in the city (Glasgow Royal Infirmary) ED clinical staff who provide clinical support. PHOENIx offers holistic, first contact, continuous primary health and social care, understanding that motivation fluctuates, and patients’ trust in care providers is usually low due to previous traumatic life experiences e.g. assault, abuse, or stigma.

Pharmacists in the PHOENIx team had undergone additional training and passed a theory and practical examination in therapeutics and clinical assessment, to qualify as independent prescribers, able to prescribe any medicine within their competency. Pharmacists carry a kit bag containing: a prescription pad; laptop for accessing clinical records; mobile phone; basic clinical equipment for common clinical assessments; medicines for emergency purposes (salbutamol, aspirin, glyceryl trinitrate spray); wound care products; lists of relevant health and social care contacts; naloxone and clean injecting equipment. They have read and write access to patients’ clinical records (addictions/mental health teams, Homeless Health Service General Practice Hospital, community pharmacy dispensing) and the third sector Homeless link worker has read and write access to wider third sector contacts. Working in pairs, the team gather as much information as possible about each patient on first meeting, using medical and third sector charity records, to understand the patient’s priorities and inform an individualised intervention and in case of any known risks. The pharmacist conducts a clinical assessment, then prescribes, refers or treats patients on the spot.

The Homeless link workers remain employed as part of their parent charity organisation, although have NHS honorary contracts. During consultations, the link worker offers to maximise the patient’s incoming financial benefits, improve access to better accommodation, encourage participation in social activities and generally problem solve where possible in any other aspect of the patient’s shared story while building a therapeutic relationship.

In addition to clinical/technical expertise, selection procedure at interview for PHOENIx staff included demonstration of empathy, non-judgemental attitude and active listening.

### Interventions

#### Usual care

In Scotland, PEH are offered a temporary single room in a designated city centre venue e.g. hotel, hostel or bed and breakfast accommodation, and allocated a named case worker.

Patients with problem alcohol or substance use may receive care and treatment from Glasgow’s Alcohol and Drug Recovery Service (ADRS) or the Homeless Addictions Team (HAT) or the Heroin Assisted Treatment service. Patients can present to any ADRS seeking help and receive same day assessment. In some circumstances following management of any immediate care needs they may be supported to engage with another ADRS closer to their temporary accommodation or to which they remain open from a previous treatment episode. If transport to a different base is required at the time they present the service will offer a taxi to facilitate this journey. For people already open to drug and alcohol services their care and treatment is provided through a combination of phone and face to face contact either at the base or on outreach, dependent on individual needs and circumstances.

To access primary health care including a General Practitioner, or ADRS, PEH must either travel to their registered mainstream or specialist homelessness General Practice or phone, which requires PEH to have access to a mobile phone (which can be supplied by ADRS or HAT) or use a landline within their accommodation, if appropriate. All mainstream services operated triage during COVID-19 lockdown, with requests for patients to phone the relevant care team prior to presenting at the premises, if possible. The Homeless Health Service GP practice offered phone inreach to a variety of homeless accommodation services and restarted outreach after a period of interruption.

For patients with mental health problems without problem drug use, access is through General Practitioner referral to general mental health services or a request for support via ADRS if currently linked in for treatment of problem substance use.

Clinical records for PEH are spread across three or four different platforms because different clinical teams (emergency care, specialist Homeless GP, mainstream GP, ADRS and mental health, social care) are distinct, each record system with different access privileges used by staff within each team. The Pharmacists in the PHOENIx team obtained permissions to remotely access all possible health and social care records on outreach, to understand all of the patients’ previous health and social care history and relieve patients of the burden of repeating their traumatic stories again, and for safety reasons.

In the UK, the out-patient (ambulatory) management of PEH with chronic diseases takes place either solely in primary care or between primary care and Hospital based out-patient clinics. During COVID-19 lockdowns most out-patient appointments were switched to phone or on line video consultations which may or may not have been possible for PEH. Patients in need of urgent hospital care self-present or may be referred by their GP or others to a hospital ED from where they may be admitted to hospital or discharged back to primary care. Prescribing is undertaken by GPs, and independent prescribers e.g. pharmacists or nurses with advanced clinical skills and knowledge. All prescriptions are obtained free of charge from community pharmacies. The capacity for outreach from services that PEH use is variable across the city.

### Intended purpose of the PHOENIx intervention

The PHOENIx intervention aims to decrease emergency service use, and overdoses, by increasing access to holistic preventative primary health care and improving the socioeconomic factors associated with homelessness e.g. income, housing. Offering weekly visits, on assertive outreach and through persistent follow up, PHOENIx aims to provide ‘whole person’ help for all health and social care needs: physical health; mental health; problem drug use; benefits; accommodation; and social prescribing.^30-33^ Access to the team for any reason, was facilitated by a dedicated phone number which patients could call anytime, or through the staff working for the Simon Community Scotland or the Marie Trust, who facilitated contacts face to face. The team do not discharge any patient and all contacts were in addition to the patient’s usual care. If the team cannot locate patients they enquire in accommodation or other venues or through contacts within homeless charities or statutory services, to help track PEH.

The team are aware of barriers to accessing care among PEH, and the problems posed by segmentation of services, so they adopted a person-centred, trauma informed approach. A comprehensive health and social care assessment is offered on the first meeting with each patient unless the patient’s priorities take the conversation in another direction, in which case the assessment is conducted in stages thereafter. If the team are unable to provide direct health/social care help immediately, they problem solved with the patient, on the spot e.g. referred, or made an appointment on the patient’s behalf e.g. to attend an appointment with mental health team while booking transport and providing reminders. The PHOENIx team treat patients as equal partners in discussions about their treatment and wishes, asking patients to prioritise what they want help with over the course of weekly visits for at least 6 months. For example, if the patient does not want to tackle their drug use, this is not prioritised by the team. Instead, other priorities e.g. mental health problems, or respiratory function, or financial benefits, or changing accommodation, became the focus until the patient was ready to address their problem drug use. The team provides a variety of supports at different times depending on the patient’s needs including advocacy, clothing, emotional support, phones, books, shopping, furniture.^31^ Patients injecting drugs will be routinely offered dry blood spot BBV testing, injecting equipment and naloxone at each visit. Pharmacy Services within NHS Greater Glasgow and Clyde will remain the Pharmacists’ employers throughout the study: strong communication links between the pharmacists and all other clinical teams involved in the care of PEH minimised any theoretical clinical risks.

The Intervention will be discontinued if the patient: loses capacity; enters a rehabilitation facility; becomes incarcerated and cannot not be reached in prison; becomes lost to follow up (some PEH flee violence and their whereabouts can suddenly change to prevent encounters with known assailants); or if the patient asks the team to stop offering the intervention.

The study Principal Investigator will visit pairs of PHOENIx staff on outreach every month and receive feedback from teams to assess fidelity of the intervention against the intervention domains. The team will meet monthly to discuss progress and patient scenarios, forming a robust peer support network, with daily communication through an encrypted social media channel approved for use by NHS Greater Glasgow and Clyde. The pharmacists will meet with the Homeless Health Service GPs monthly to discuss difficult cases and seek advice when necessary from the Homeless Health GP service and consultants in the ED of the local hospital and homeless liaison nurses in the local hospital. There will be regular communication with ADRS/HAT staff.

All other interventions and concomitant care are permitted throughout the trial.

### Outcomes

The co-primary endpoint is whether to progress to a definitive trial, based on any improvement in the rate of presentation to EDs for overdoses or other causes, during the 6 or 9 month follow up period, and achievement of the following progression criteria:

- recruitment of at least 100 patients within 4 months;
- ≥ 60% patients remaining in the study at 6 and 9 months follow up (excluding those who have died or lost capacity);
- establishment of the pharmacist intervention (≥ 60% of patients in the intervention group receiving the intervention as planned excluding those who have died or lost capacity);
- ≥ 80% of patients with data collected as planned (excluding those who have died or lost capacity).

Secondary outcomes are as outlined below, compared between Intervention and Usual care groups at 6 and 9 month follow up:

1. Health care utilisation which includes number of (and number of patients with):
  - Non prescribed drug overdoses;
  - hospital admissions;
  - prescribing for multimorbidy (proportion of patients prescribed medicines for diagnosed conditions; proportion of patients with minimum doses of medicines for diagnosed conditions);
  - contacts (phone or face to face) with GP/nurse/addictions worker/other healthcare professional;
  - Scottish Ambulance Service call outs;
  - missed out-patient appointments.
2. Time from randomisation until: first ED visit for OD and other reasons; death; and hospitalisation.
3. Patient reported measures:
  - EuroQol 5D 5L quality of life score ^34^
  - Patient Experience with Treatment and Self-management measure (PETS)^35^
  - Frailty score ^36^
  - Anxiety/depression ratings
  - Modified MRC breathlessness scale

Outcomes will be collected by independent researchers interviewing patients and accessing NHS clinical records at 6 and 9 month follow up compared between the intervention and usual care groups. Intervention acceptability will also be measured through a parallel qualitative evaluation.

### Participant timeline

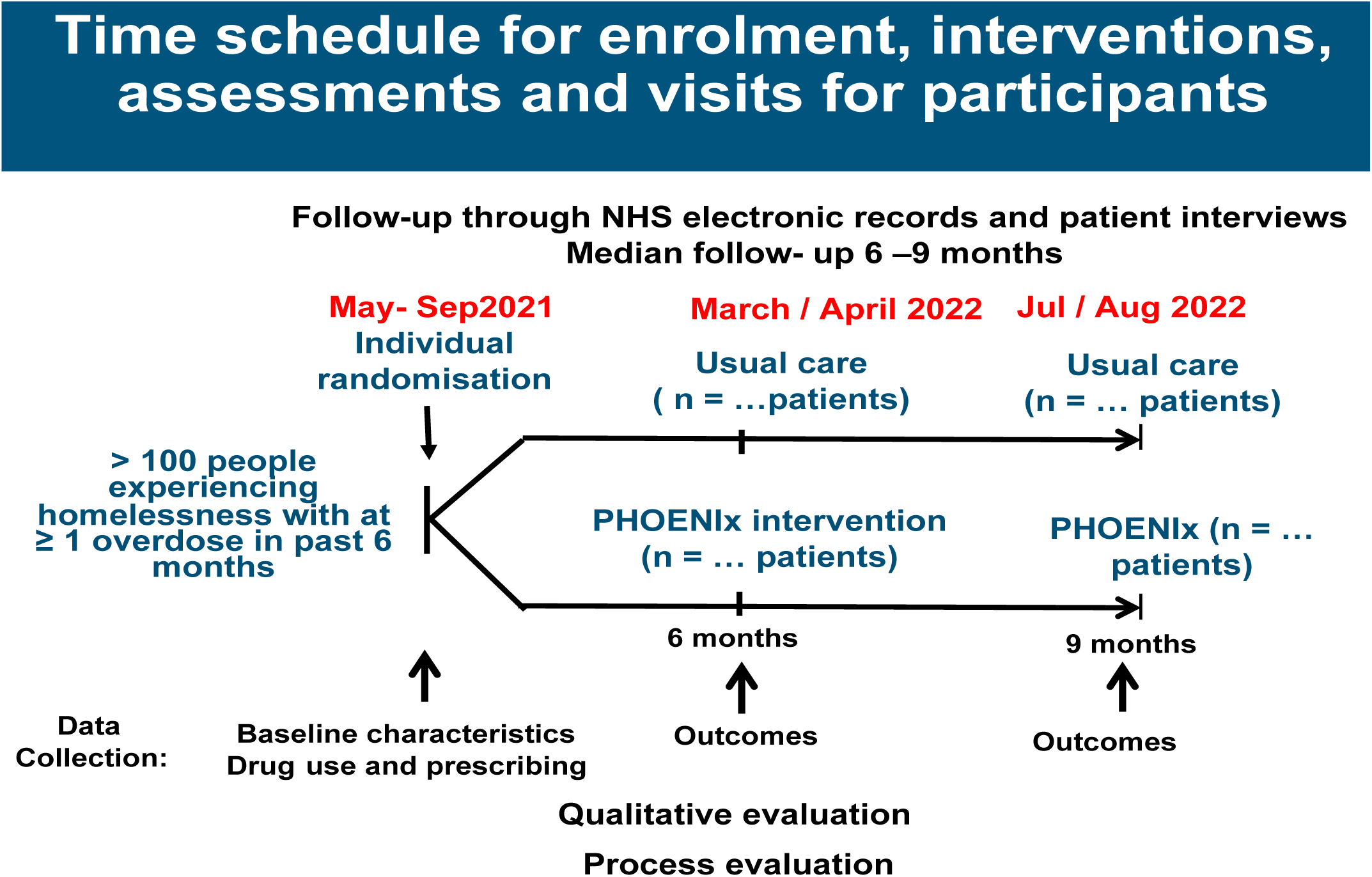

### Sample size

The guidance on sample sizes for pilot studies varies with 30-50 patients per arm thought to be sufficient, because the focus is on estimating parameters for the full study, rather than formal testing of hypotheses.^37^ We aimed to invite approximately 160 patients, anticipating a recruitment rate of ∼60% based on our earlier feasibility study.^32^ If 100 agree to participate, we estimate the recruitment rate as 62.5% (95% CI 55%-70%). Mortality rate in our previous feasibility study was 8.3% over one year, therefore we anticipated 6 patients dying over 9 months.^32^ Assuming a conservative retention rate of 70% after 9 months, and additional losses due to mortality, we anticipate at least 64 patients with 6 and 9 month follow up data to inform sample size for a full scale randomised controlled trial.

### Recruitment

#### The importance of researcher ‘fit’

Induction training will include visits to potential study sites in Glasgow, and introductions to accommodation staff and third sector street homeless outreach workers were in different venues. Processes for obtaining consent and baseline and follow up interviews and data collection will also be explained and rehearsed. The researchers comprise a qualified mental health nurse with more than 20 years of experience in front line NHS service delivery and research with PEH, and those experiencing problem substance use and poor mental health; and a pharmacy technician with over 20 years’ experience in patient services (mental health and general hospitals).

### Process

Researchers will visit accommodation and other venues, to approach all potentially eligible patients, face to face. Patient self-report of overdose will be confirmed by examination of clinical records and/or testimony from witnesses e.g. accommodation staff, friends, or injecting/drug using partners.

Confirmation included ambulance call out, naloxone administration recorded in clinical notes or in-house patient records made by accommodation providers, or an ED visit for overdose. Researchers will ask each potential recruit about the circumstances of the overdose, and when this occurred, including whether the patient had any recollection of having received any assistance from other people at the time of overdose. This approach to identifying eligible patients will be taken because our collective clinical experience suggested most non-fatal overdoses are not formally recorded or if recorded, there is no standardised, identifiable coding applicable across different clinical / administrative records. Patients will be offered a non cash incentive (voucher for use in a city centre store not selling alcohol or tobacco) of £10 (equivalent to 13 US dollars or 12 Euros) on completion of baseline data collection, and after completion of each follow up data collection at months 6 and 9. The baseline interview pro-forma is shown in Supplementary information.

### Methods: assignment of interventions

#### Allocation: sequence generation

One hundred and sixty sealed opaque envelopes will be generated remotely by staff from the University of Birmingham not directly involved in participant recruitment. Each envelope will contain a folded piece of paper with the computer generated printed words: ‘PHOENIx Intervention’ or ‘Usual care’. The envelopes will be randomly shuffled, by staff from the University of Birmingham then sent in a box, by secure mail, to the Glasgow study centre before the first patient is recruited. This is an individual level randomisation approach without stratification.

#### Allocation concealment mechanism and implementation

Researchers will take informed consent by discussing the patient information leaflet with patients, explaining what the study entails, and asking if the patient would want to participate. Some patients will read the information and make the decision themselves. In both cases, patients will have time to read the information or have it explained to them, and ask questions before coming to a decision. Researchers will conduct baseline interviews in person with participants in different venues in Glasgow city centre. At the end of the interview researchers will phone the study centre asking for a randomisation. One of the research team will answer the call immediately and, in the presence of another member of the research team, pick an envelope at random from the box of envelopes. The research team will request the following information from the researcher: patient name; date of birth and location. A sequential study number will be written on the outside of the chosen envelope and, in the presence of another member of the research team, and while the researcher remains on the phone, the envelope will be opened and the allocation revealed to the researcher (and patient) on the phone after two members of the research team read the allocation from the piece of paper inside the envelope. The study team will return the piece of paper inside the envelope then write the words ‘Intervention’ or ‘Usual care’ onto the outside of the envelope together with the date and store the envelope inside a folder (one folder per patient), inside a lockable filing cabinet within the study centre. Participants will therefore be randomised in a one-to-one ratio to ‘usual care’ or ‘PHOENIx intervention’. On allocation to Intervention, the patient’s details and location will be communicated to the PHOENIx team who will be asked to contact the patient and begin offering the intervention.

Following allocation to usual care, participants will have no further contact from study personnel until follow up data collection.

#### Blinding

Independent statisticians conducting analysis of follow up data will be blinded to allocation. Assessment of outcomes from clinical records will be conducted by a researcher / administrator who will be blind to assignment to intervention or usual care group.

### Data collection, management and analysis

Baseline, six and nine month follow up data will be collected during researcher led face to face patient interviews in the patient’s accommodation, or in homeless charity drop in centres in Glasgow city centre. Interviews will last approximately 45 minutes. Study instruments used during interviews e.g. weighing scales, peak flow meters, were used routinely during clinical care in primary and secondary care, and familiar to researchers.

Data to be collected (Supplementary information) include: details of comorbidities; non prescribed drug use; prescribed medicines; health related quality of life using EQ-5D-5L; weight; height; Blood pressure; Pulse; Peak Expiratory Flow rate; modified Medical Research Council Dyspnoea scale; COPD Assessment Test; grip strength, GP visits, nurse visits, ADRS and mental health team and other primary healthcare contacts; un/scheduled secondary healthcare contacts including missed out patient appointments; Fried’s Adapted frailty phenotype questions and an adapted version of the Patient Experience with Treatment and Self-Management (PETS) measure.

The intervention team will record all treatment administered e.g. dressings changes, prescriptions issued, referrals; advice given; gifts and kindness e.g. personal support in form of clothing or food; advocacy and type (face to face or phone call) of patient visits.

Validated questionnaires used during interviews have not previously been used in PEH, therefore, the research and clinical team evaluated their suitability in advance and decided only one needed modification: The PETS.^35^ Some aspects of the language used in the PETS and assumptions about how patients obtained their medicines and aspects of self-care were deemed potentially incomprehensible to PEH in Scotland, which led to the research team making minor modifications, which were discussed with the questionnaire author. They included, for example, replacing the term “refill” in relation to medicines, to “get more of your medicines before they run out” or replacing “somewhat” with “neither easy nor difficult” or “monitor” with “keep tabs on” or “checking blood pressure” with “checking injection sites”. The section containing five questions about “Medical and healthcare expenses” was omitted because the health service in Scotland does not charge for care and all prescriptions are free.

Interview data will be recorded on paper data collection forms during interviews, prior to transcription onto an EXCEL spreadsheet by the research team. Prescribing, co-morbidities, laboratory test values, General Practitioner contacts and other healthcare utilisation data will be subsequently extracted from medical and ADRS team records and entered onto the same EXCEL spreadsheet. We therefore plan to utilise data from these two sources (patient reports and data from medical records) to provide a comprehensive picture. Members of the research team will cross check a 10% sample of data entries for accuracy and completeness. At 6 and 9 month follow up, the research team will make repeated attempts to re-engage patients, as will the PHOENIx team during the intervention phase. Repeated attempts will be made weekly and in addition to phone calls (if the patient has a phone) including: checking primary care clinical records for attendance at health and social care venues for appointments and meeting the patient at the appointment; checking emergency department and hospital records for presentations and changes to contact address and contact numbers or for change of status from having capacity to loss of capacity (reason for withdrawal from the trial) or death; phone/visit to community pharmacies to ask if the patient has attended for collection of prescription or for supervised opiate substitution treatment; phone/visit to the patient’s accommodation to enquire about the patient’s usual patterns of appearance; asking the homeless community hub staff if patient has attended; asking the third sector homeless outreach teams for possible sightings; and visiting the patient’s usual begging site. If patients cannot not be located, researchers will still be able to collect patient data from hospital records, General Practices and Alcohol and Drug Recovery Services as appropriate. If patients die or loose capacity, data up until the point of death or loss of capacity, will be collected.

### Statistical methods

Outcome analysis will be conducted by independent statisticians at University of Birmingham after collection of 9 month follow-up data. Primary outcome measures will be described using proportions, along with 95% confidence intervals to describe uncertainty. Patient questionnaire and clinical measures will be analysed according to the intention to treat principles. Appropriate summary statistics (e.g. proportions & inter-quartile ranges, means and standard deviations) along with 95% confidence intervals will be generated for the study feasibility and patient reported / clinical / health utilisation measures. By design there is no a-priori powered endpoint, however hypothesis testing will be conducted to determine whether there is any difference between outcome measures. Between-group measures (mean differences and relative risks) will be reported with 95% confidence intervals.

### Economic evaluation

An embedded economic evaluation will examine the feasibility of determining the cost-effectiveness of the PHOENIx intervention in a subsequent definitive trial. The main analysis will consider a health and social care service perspective whereby unit costs are applied to each item of health (e.g. hospitalisation) and social care service use data. Unit costs will be taken from routine sources where possible including missed appointments. ^38-40^ The effectiveness of the intervention will be explored in terms of health state utilities (for a future cost utility analysis), as measured using the EQ-5D-5L to generate Quality Adjusted Life Years (QALYs) to be used alongside the cost data to give an indicative picture of cost-effectiveness. QALYs will be generated from the EQ-5D-5L using appropriate crosswalk methods and applying reference values for the EQ-5D-3L.^41-43^ Both cost and utility outcomes will be quantified to describe the costs of the services and to provide QALY data, as currently few data on the QALY loss associated with homelessness are available.

### Qualitative evaluation

In a parallel process evaluation, we will explore participant perspectives of their drug use and overdoses, including aspects of support perceived as most important in order to prevent subsequent drug overdose and their perceptions of the existing pathway for health and social care follow up post drug overdose together with their experience of the intervention. This will be conducted through qualitative face-to-face semi structured interviews with a purposive sample of 20-30 recruited patients in the intervention and usual care group in order to obtain a variety of experiences. Interviews will be conducted by an independent researcher (NF) who has no knowledge of patients prior to the interviews. All interviews will be conducted in a city centre drop in service utilised by PEH. Data will be gathered by recording face to face semi-structured interviews via an audio digital recording device and will be transcribed, using pseudonyms to ensure confidentiality and anonymity. All study participants will receive a £10 voucher as recognition for their participation. Thematic coding will be conducted by NF and then checked by members of the research team to reduce the risk of bias, ensure consistency and rigour. Normalisation Process Theory (NPT) will be used to inform conceptualisation of the process evaluation data because it is a theoretical framework used to aid development, evaluation and implementation of complex interventions. NPT is a theory that focuses on the “workability” of complex interventions in the real world.^44^ We hypothesise that PEH may be “overwhelmed” by self-management tasks and will vary in their capacity to cope with any given level of treatment burden depending on a range of factors such as health literacy, language, drug seeking behaviour, level of educational attainment, personal beliefs, physical and mental abilities, and structural and practical barriers to accessing care. We will be looking at the issue of treatment burden with participants through a NPT lens and considering how the intervention affects individual capacity. Findings will be considered alongside quantitative outcomes (PETS, treatment burden measure) obtained as part of the secondary outcome measures.

This qualitative work will enable us to capture rich, complex data and unanticipated insights. Data will be analysed using NVivo V.12 software.^45^

### Data monitoring

A multidisciplinary data monitoring committee involving researchers, NHS administrators and clinicians will have oversight of the qualitative and quantitative data collection process, and study methods, independent from the main study funder (Drug Death Task Force of the Scottish Government). The data monitoring committee will ensure researchers have access to relevant data, and study milestones are adhered to. No interim analyses are planned, and as the study intervention is offered in addition to usual care, with PHOENIx supporting patients using guideline based care only, adverse events of the trial intervention are not anticipated. Trial conduct will be audited by NHS Greater Glasgow and Clyde Research and Development, independent from the study investigators.

### Ethical approval and trial registration

The trial is registered with the UK Clinical Trials Registry (ISRCTN 10585019), and was approved by the South East Scotland Research Ethics Committee 01 and subsequently, governance approval by NHS Greater Glasgow and Clyde Research and Innovation. The authors confirm that all ongoing and related trials for this intervention are registered.

## Discussion

Homelessness is a growing global public health problem associated with many adverse health outcomes, yet research in this sphere, particularly empirical evidence of multiple health, vulnerability factors, social care problems, and RCT evidence for collaborative health and social care interventions, remains sparse.^2^

The pilot PHOENIx after overdose RCT aims to determine whether a subsequent definitive RCT of a Pharmacist and third sector homeless outreach worker intervention is merited. The basis for this is achievement of progression criteria comprising recruitment, retention, data collection, and establishment of the pharmacist intervention together with any signal of reduction in emergency department presentations for overdoses or other causes.

Strengths of this pilot trial relate to its rigorous design including embedded qualitative process and economic evaluations. The intervention is complex, and phases of complex intervention testing are being followed. The intervention is aligned with best practice recommendations in relation to assertive outreach, integrated health and social care support, providing longer appointments to enable patients to discuss their multiple issues. The extensive characterization of the patient population, recruited from 20 different locations in Scotland’s largest city, ensures a cross section of PEH. Follow up is longer than most previous RCTs in homelessness. Outcomes are diverse, covering progression criteria for a subsequent randomised controlled trial, health care utilization, and objective and subjective health measures. However, limitations relate to the fact that this pilot trial is being conducted in Scotland only, with a population of PEH that may not be ethnically diverse. High levels of refusal to study participation and dropout leading to low numbers at 6 and 9 month follow up could also potentially threaten the generalizability of study results but we will be monitoring and reporting on this.

The PHOENIx pilot randomised controlled trial will provide new data and if objectives are met, seeks to inform a subsequent definitive trial aiming to reduce ED presentations for non-prescription drug overdose and other reasons, by strengthening primary health and social care in the community. It will also provide new knowledge about the characteristics of this patient population in terms of: the extent, nature of and treatment uptake for multimorbidity; non-prescribed drug use; uptake of primary and secondary health and social care; welfare benefits; treatment burden; subjective and objective measures of health including quality of life; laboratory results; and the prevalence of frailty to help inform future clinical trials and service delivery in this area.

## Data Availability

All relevant data for the study will be made available on study completion

## Access to data

The principal investigator, researchers and independent statisticians will have access to the final trial dataset.

## Dissemination policy

Trial results will first be communicated to participants individually, verbally or in writing, through existing homelessness networks and accommodation providers. Healthcare, social care and third sector workers involved in the trial (GPs, ED staff, addictions staff and others) will receive a written summary of findings with an offer to meet the Principal Investigator to discuss the results. Health and Social care / Homelessness staff with a role in policymaking will receive a summary of results, and an offer of meeting to discuss any implications. The study findings will be described by all of the research team in accordance with guidelines for eligibility of authorship and submitted for publishing in a peer reviewed journal. There are no plans to involve professional writers.

## End of study date

Processes for NHS research governance approvals were delayed during COVID-19 lockdown, leading to a delay in the trial start date. The end-of-study date is on the last day of 9 month follow up data collection (July 2022). Allowing time for data input to the trial database, summary and analysis, the final results will be available in the last quarter of 2022.

## Conclusion

We expect this pilot RCT aimed at PEH with recent non–prescription drug overdose will determine whether a full RCT is justified and to provide valuable insights into the health and social care needs of this marginalised population, while providing a signal of whether the complex PHOENIx health and social care intervention improves outcomes.

